# Antimicrobial Resistance pattern in Gram-Negative Uropathogens in Duhok City, Northern Iraq

**DOI:** 10.1101/2023.02.23.23285568

**Authors:** Razvan Luqman Yasen

## Abstract

Antimicrobial resistance (AMR) is one of the most dangerous global threats since antimicrobial discovery. The world health organization(WHO) has implemented a program called GLASS to mitigate resistance across the globe. Urinary tract infection(UTI) are the second most common infections and are the most common reason for prescription of antimicrobials, the rise in AMR has caused concerns of UTI Overuse and misuse of prescriptions and decrease of treatment options hence many researches conducted across the globe are on uropathogens resistance rate and trend. This retrospective study was conducted in duhok province of KRI to measure antimicrobial resistance percentages and identify the most common uropathogens.

309 urine samples were collected in a time span of 12 months. Urine samples were collected by clean catch midstream and inoculated on blood and MacConkey agars, Antibiotic sensitivity test (AST) was performed to identify Gram negative uropathogen and its sensitivity pattern.

We found out most common Gram negative uropathogen in females were E.coli and Klebsiella pneumonia while in males it was E.coli and Pseudomonas aeruginosa and common Klebsiella pneumonia. E.coli was most resistance to amoxicillin/amp(64.2%) and it was least resistant to carbapenems(6.1%). Klebsiella pneumonia had similar resistant pattern to E.coli. pseudomonas aeruginosa was highly resistant to all antimicrobials, third gen cephalosporins were the highest 95.7%.

AMR has risen to concerning levels in duhok and if not controlled would result in simple infections causing death in future we recommend guidelines for control of Overuse, misuse and ease of availability of antimicrobials as a measure to decrease AMR. Continues monitoring should be performed on AMR development in the future.

## Introduction

The World Health Organization (WHO) has designated antimicrobial resistance (AMR) as one of the major global threats, therefore the WHO took the necessary steps to mitigate the impact of AMR on global health (1). AMR caused increased mortality and morbidity, prolonged hospital stays, usage of more expensive with more serious side effects of second and third-line treatments (2). Additionally new medical advances like joint replacement, organ transplants, cancer therapy and treatments of chronic diseases rely on Antimicrobials (AM) to suppress infections (3, 4). Statistically 2 million people in the USA and 400,000 thousand people in Europe are infected yearly by AMR with 23,000 and 25,000 dying due to infection, respectively (4). AMR can develop and spread through humans, animals, agriculture, and environment, one of the most important reasons of AMR occurrence is due to AM excessive prescription and self-medication (which is worse in developing countries due to ease of availability) inappropriate drug choice, wrong dosing, not adhering to treatment instructions (4-7). AMR are categorized into 4 mechanisms, Limiting Drug intake through an outer membrane which only gram negative Species possess; Modifying Drug target, AM’s can target a variety of bacteria components which the bacteria will try to modify to enable resistance; drug inactivation either through drug degradation or transferring of a molecule to the drug to inactivate it; drug efflux through efflux pumps which get rid of toxic components inside bacteria these pumps can transport a variety of compounds which result in Multi-drug efflux pump (8, 9).

Urinary Tract Infections (UTI) can be upper or lower UTI classified as pyelonephritis or cystitis, respectively (10, 11). UTI are the second most common infections after respiratory tract infections. Estimation put UTI cases annually at 150 million cases per year [6]. The most UTI cases affect women because of shorter distance to bladder and urethral Opening closer to Rectum (12). According to statistics more than 50% of women will suffer UTI’s during their lifetime (13). UTI Sings and symptoms are urgency, dysuria, suprapubic tenderness, fever, chills nausea, vomiting (14). physicians often prescribe antibiotic therapy before identifying bacterial organism because of characteristic and worrying symptoms that need immediate treatment (15).

Causative UTI Bacteria species are classified into Gram-positive and Gram-Negative organisms, with the Gram-Negative dominating UTI cases about 90% of the time. Escherichia coli alone accounts for 80% of gram-Negative UTI cases (16), Klebsiella Pneumonia makes up the rest 5-20% of Gram-Negative cases (17), gram positive cases are less common and include mostly staphylococcus saprophyticus, enterococcus spp. And Streptococcus group B (17).

Every Region must do a local survey of bacterial prevalence and susceptibility to make empirical guidelines, to make sure physician prescribe the right Empirical antibiotic which will reduce AMR drastically as misuse of prescription is the biggest cause of resistance. Previously, antibiotic sensitivity patters were studied in Iraq (5, 18-23). It was found that multidrug resistant microorganisms are common in the Iraq (9, 24-28). Our aim in this study is to collect and analyze 10 months’ worth of UTI cases in Duhok city Kurdistan Region of Iraq.

## Materials and Methods

### Study Setting

this retrospective cross-sectional study was conducted in Duhok city, Kurdistan region of Iraq (KRG) by retrieving recorded data from 2010-2022 between September, 2022 to March 2023. The data was collected from Azadi tertiary hospital which serves 1.5 million people in Duhok province located west of KRG and surrounding area.

### Sample Collection and Antibiotic Sensitivity Test

The data was recorded by performing Antibiotic Sensitive Test (AST) for urine samples of patients attending Azadi hospital, the urine samples were collected by clean catch midstream which afterwards were inoculated on blood and MacConkey agars. AST was done to identify the bacterial organism and its antimicrobial sensitivity, which was performed either Manually or Automated (VITEK) there was no discrimination made between the two methods of data collection. The recorded data collected had information on the following: name, gender, bacterial organism, and bacterial sensitivity and resistance.

### Ethics

The Study protocol was approved by the ethics committee of the college of medicine, University of Duhok.

## Results

### Patient characteristics

In this retrospective study a total of 309 cases were collected from duhok province, 211(68.3%) of the cases were female and 97(31.4%) were of male cases.

### Prevalence of UTI infections

According to Table 1 The most common and predominant UTI gram-negative bacteria were E.COLI with 218(70.55%) cases followed by 43(13.9%) cases of Klebsiella pneumonia. The less common bacteria were Pseudomonas aeruginosa with 23(7.44%) and Proteus with 7(2.26%) cases.

**Table 1:**
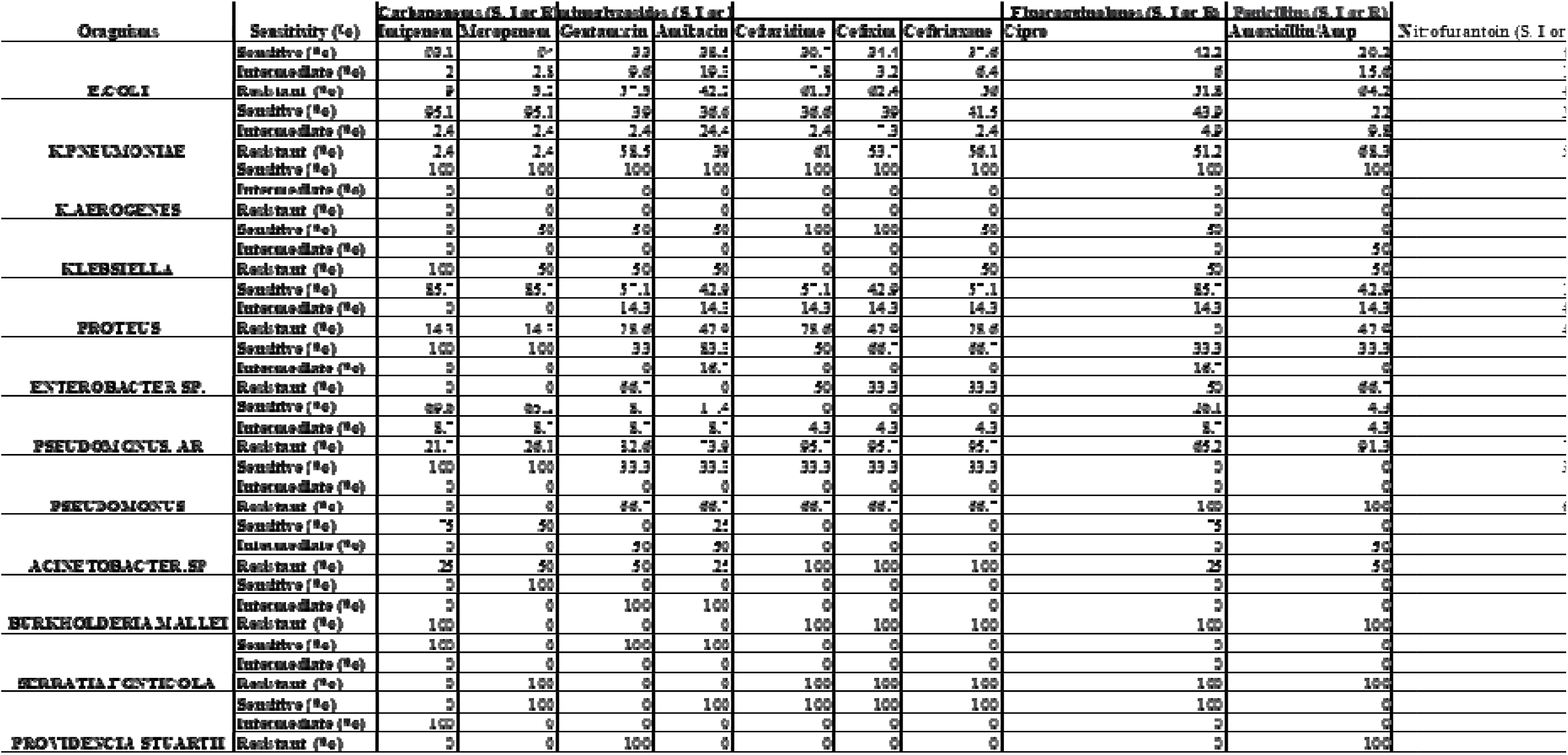
Representing total antibiotic sensitivity results.

**Table 2:** Representing female only sensitivity pattern (N/A means no cases)

| Oragnisms | Sensitivity (%) | Carbapenems (S, I or R) |  | Aminoglycosides (S, I or R) |  |  |  |  | Fluoroquinolones (S, I or R) | Penicillins (S, I or R) | Nitrofurantoin (S, I or R) |
| --- | --- | --- | --- | --- | --- | --- | --- | --- | --- | --- | --- |
|  |  | Imipenem | Meropenem | Gentamicin | Amikacin | Ceftazidime | Cefixim | Ceftriaxone | Cipro | Amoxicillin/Amp |  |
| E coli | Sensitive (%) | 94.3 | 94.3 | 32.5 | 38.9 | 36.9 | 42 | 44.6 | 47.8 | 21.7 | 50.3 |
|  | Intermediate (%) | 0 | 2.5 | 10.2 | 20.4 | 9.6 | 3.8 | 7.6 | 5.1 | 17.8 | 12.1 |
|  | Resistant (%) | 5.7 | 3.2 | 57.3 | 40.8 | 53.5 | 54.1 | 47.8 | 47.1 | 60.5 | 37.6 |
| k pneumoniae | Sensitive (%) | 96.8 | 96.8 | 48.4 | 41.9 | 45.2 | 48.4 | 48.4 | 54.8 | 25.8 | 32.3 |
|  | Intermediate (%) | 3.2 | 3.2 | 3.2 | 16.1 | 3.2 | 6.5 | 3.2 | 6.5 | 9.7 | 22.6 |
|  | Resistant (%) | 0 | 0 | 48.4 | 41.9 | 51.6 | 45.2 | 48.4 | 38.7 | 64.5 | 45.2 |
| k.aerogenes | Sensitive (%) | 100 | 100 | 100 | 100 | 100 | 100 | 100 | 100 | 100 | 0 |
|  | Intermediate (%) | 0 | 0 | 0 | 0 | 0 | 0 | 0 | 0 | 0 | 100 |
|  | Resistant (%) | 0 | 0 | 0 | 0 | 0 | 0 | 0 | 0 | 0 | 0 |
| KLEBSIELLA | Sensitive (%) | 0 | 0 | 0 | 0 | 100 | 100 | 100 | 0 | 0 | 0 |
|  | Intermediate (%) | 0 | 0 | 0 | 0 | 0 | 0 | 0 | 0 | 100 | 100 |
|  | Resistant (%) | 100 | 100 | 100 | 100 | 0 | 0 | 0 | 100 | 0 | 0 |
| PROEUS | Sensitive (%) | 83.3 | 83.3 | 66.7 | 50 | 50 | 50 | 50 | 83.3 | 33.3 | 16.7 |
|  | Intermediate (%) | 0 | 0 | 0 | 16.7 | 16.7 | 0 | 16.7 | 16.7 | 16.7 | 33.3 |
|  | Resistant (%) | 16.7 | 16.7 | 33.3 | 33.3 | 33.3 | 50 | 33.3 | 0 | 50 | 50 |
| ENTEROBACTER SP. | Sensitive (%) | 100 | 100 | 66.7 | 100 | 66.7 | 100 | 100 | 66.7 | 33.3 | 33.3 |
|  | Intermediate (%) | 0 | 0 | 0 | 0 | 0 | 0 | 0 | 0 | 0 | 0 |
|  | Resistant (%) | 0 | 0 | 33.3 | 0 | 33.3 | 0 | 0 | 33.3 | 66.7 | 66.7 |
| PSEUDOMONUS. AR | Sensitive (%) | 66.7 | 66.7 | 11.1 | 22.2 | 0 | 0 | 0 | 22.2 | 0 | 0 |
|  | Intermediate (%) | 0 | 0 | 11.1 | 11.1 | 11.1 | 11.1 | 11.1 | 11.1 | 11.1 | 22.2 |
|  | Resistant (%) | 33.3 | 33.3 | 77.8 | 66.7 | 88.9 | 88.9 | 88.9 | 66.7 | 88.9 | 77.8 |
| PSEUDOMONUS | Sensitive (%) | 100 | 100 | 100 | 100 | 100 | 100 | 0 | 0 | 0 | 100 |
|  | Intermediate (%) | 0 | 0 | 0 | 0 | 0 | 0 | 0 | 0 | 0 | 0 |
|  | Resistant (%) | 0 | 0 | 0 | 0 | 0 | 0 | 100 | 100 | 100 | 0 |
| ACINETOBACTER.SP | Sensitive (%) | N/A | N/A | N/A | N/A | N/A | N/A | N/A | N/A | N/A | N/A |
|  | Intermediate (%) | N/A | N/A | N/A | N/A | N/A | N/A | N/A | N/A | N/A | N/A |
|  | Resistant (%) | N/A | N/A | N/A | N/A | N/A | N/A | N/A | N/A | N/A | N/A |
| BURKHOLDERIA.MALLEI | Sensitive (%) | N/A | N/A | N/A | N/A | N/A | N/A | N/A | N/A | N/A | N/A |
|  | Intermediate (%) | N/A | N/A | N/A | N/A | N/A | N/A | N/A | N/A | N/A | N/A |
|  | Resistant (%) | N/A | N/A | N/A | N/A | N/A | N/A | N/A | N/A | N/A | N/A |
| SERRATIA.FONTICOLA | Sensitive (%) | 100 | 0 | 100 | 100 | 0 | 0 | 0 | 0 | 0 | 0 |
|  | Intermediate (%) | 0 | 0 | 0 | 0 | 0 | 0 | 0 | 0 | 0 | 0 |
|  | Resistant (%) | 0 | 100 | 0 | 0 | 100 | 100 | 100 | 100 | 100 | 100 |
| providencia stuartii | Sensitive (%) | 0 | 100 | 0 | 100 | 100 | 100 | 100 | 100 | 0 | 0 |
|  | Intermediate (%) | 100 | 0 | 0 | 0 | 0 | 0 | 0 | 0 | 0 | 0 |
|  | Resistant (%) | 0 | 0 | 100 | 0 | 0 | 0 | 0 | 0 | 100 | 100 |

**Table 3:**
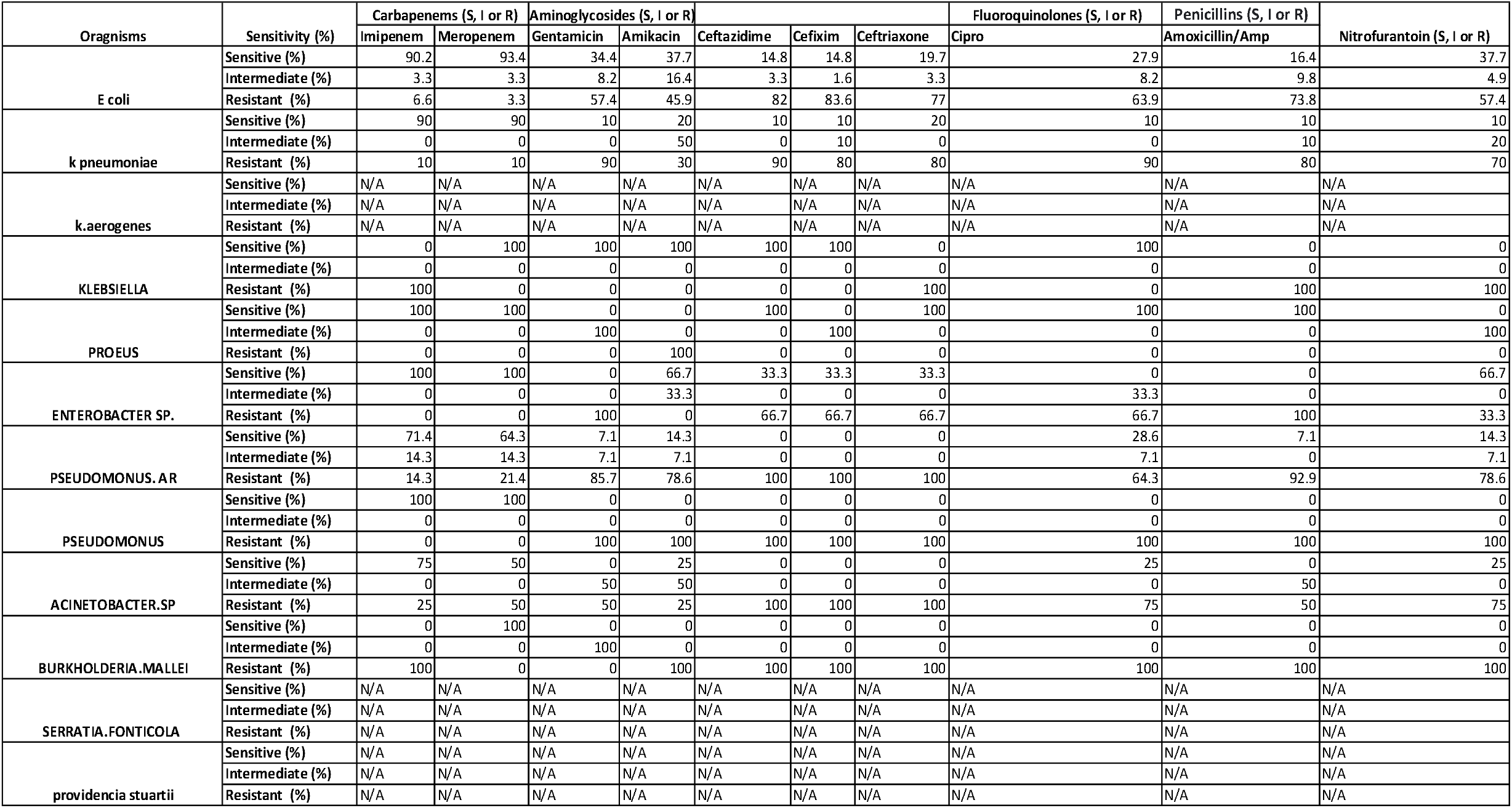
Representing male only sensitivity pattern (N/A means no cases)

E.coli highest reported resistance was against the penicillin drug amoxicillin/amp 64.2% followed closely by cephalosporins third generation drugs cefixime 62.4%, ceftazidime 61.5%, ceftriaxone 56%, aminoglycosides drugs gentamycin 57.3% and amikacin 42.2%, fluoroquinolone ciprofloxacin was 51.8%, nitrofurantoin was 43.1%. E.coli had the lowest resistance against carbapenems 3.2% meropenem, 9% imipenem.

K.Pneumonia resistance pattern was similar to E.coli with a 0.5-4% difference in each drug resistance, amoxicillin/amp was the most resistance 68.3% and carbapenems least resistant imipenem and meropenem 2.4%, the exceptions to these similarities were cefixime 53.7% and imipenem 2.4% which had 7.65% less resistance and nitrofurantoin 51.2%, 8.1% more resistance.

While Pseudomonas aeruginosa were the less common gram-negative bacteria according to our data it has the highest resistance to all drugs, all three third-gen cephalosporin drugs were 95.7% making them the most resistance antibiotics(ceftazidime, cefixime and ceftriaxone) followed by amoxicillin/amp 91.3%, gentamicin, nitrofurantoin, amikacin, Cipro were 82.6%,78.3%, 73.9%, 65.2%, respectively, the least resistance drugs but still high were carbapenems imipenem and meropenem with 21.7%, 26.1, respectively.

### UTI Infections rate among females

In females E.coli is the predominant UTI gram-negative bacteria with 166(78.67%) cases followed by Klebsiella pneumoniae 31(14.69%) cases less common were pseudomonas aeruginosa 9(4.27%) and PROTEUS 6(2.84%) cases.

E.coli in females if compared to male E.coli cases had lower resistance to all drugs, with amoxicillin/amp reported highest resistance 60.5% followed by gentamicin 57.3%, cephalosporins cefixime, ceftazidime, ceftriaxone made up 54.1%, 53.5%, 48.4%, respectively, ciprofloxacin, amikacin, nitrofurantoin were 47.1%, 40.8%, 37.6% respectively, the lowest resistant drugs were the carbapenems meropenem and imipenem 3.2%. 5.7%, respectively.

Klebsiella pneumonia had highest resistance to amoxicillin/amp 64.5% and least resistance to carbapenems reported 0% resistance.

Pseudomonas aeruginosa was uncommon but had the highest resistance reported to drugs, highest was 88.9% to gen three cephalosporins and amoxicillin/amp, 77.8% amikacin and nitrofurantoin, 66.7% to ciprofloxacin, and least amount of resistance to carbapenems 33.3%.

### UTI Infection rate among male males

E.coli made up most of male UTI bacteria cases 61(62.9%) followed by pseudomonas aeruginosa 14(14.4%) and Klebsiella pneumonia 10(10.3%) cases, less common was Acinetobacter spp. with 4(4.1%).

E.coli in males had most resistance to cephalosporins cefixime, ceftazidime, ceftriaxone with 83.6%, 82%, 77%, Respectively, followed by amoxicillin/amp with 73.8%, ciprofloxacin was reported 63.9%, gentamicin and nitrofurantoin both had 57.4%, amikacin was 45.9%, the least resistance drugs were carbapenems meropenem, imipenem with 3.3%, 6.6%, Respectively.

Pseudomonas aeruginosa is not only the second most common among men but also most dangerous, its resistance rate is the highest among the other bacteria 100% reported resistance against all three cephalosporins, 92,9% against amoxicillin/amp, 85.7% gentamicin, amikacin and nitrofurantoin both have 78.6% resistance, ciprofloxacin was 64.3%, the least resistant drugs were carbapenems imipenem and meropenem making up 14.3%, 21.4%, Respectively.

Klebsiella pneumonia among men has high resistance according to our data the most resistant are gentamicin, ceftazidime, ciprofloxacin with 90%, followed by amoxicillin/amp, cefixime, ceftriaxone 80%, nitrofurantoin was reported 70%, the least resistant were amikacin 30% with the carbapenems imipenem and meropenem making up the lowest resistant 10%.

## Discussion

Estimations have put mortality cause of bacteria antimicrobial resistance in 2019 at 4.95 million associated with AMR with 1.27 million contributed by it, six of the leading bacteria’s 4 of which are included in this study have led to worldwide deaths of 0.929/1.27 million people and associated deaths of 3.57/4.95 million people, E.coli was responsible for most deaths followed by Klebsiella pneumoniae (29, 30). AMR is a global problem and needs to be dealt with on a national basis hence this study was conducted to assess the resistant pattern of the most common gram-negative UTI bacteria which will paint a picture of the AMR situation in duhok province.

Females (68.3%) were more likely to be infected with UTI than males (31.4%) in our study similar results were found in other studies(31), in females the most common UTI bacteria was E.coli(78.67%) followed by klebsiella pneumoniae(14.69%) these finding were in agreement with (14, 31, 32), while in males E.coli(62.9%) followed by pseudomonas aeruginosa(14.4%) and klebsiella pneumoniae(10.3%) were the most common these results were in agreement with (33). the results might be lower in these researches because they count gram negative and positive as combined.

E.coli was most resistant to penicillins(64.2%) followed by gen three cephalosporins(59.96%) and gentamicin(57.3%), ciprofloxacin(43.9%), nitrofurantoin(43.1%), amikacin(42.2%), the least resistant antibiotics were carbapenems(6.1%) similar results were found in(34, 35) and also in (32, 33) although amikacin and nitrofurantoin were low in these two studies reasons for this could be due to low cases of these two antibiotics in these studies.

In our data e.coli had higher resistance for most antibiotics in males than females, AMR develops in the same fashion in both genders these difference might be due to cultural reasons, ease of availability of drugs, drug abuse by males, or females more adherent to physicians prescriptions.

Klebsiella pneumoniae had similar resistance pattern to e.coli these findings were also mentioned in (36) albeit results from other researches were very different from our results(32, 33, 36) besides carbapenems ceftriaxone, ciprofloxacin, nitrofloxacin resistance percentage these differences might be due to different methodology and lower cases of klebsiella pneumonia in these studies. the similarities between e.coli and klebsiella pneumonia resistance pattern could be due to them both being in the Enterobacteriaceae family, both are found in similar environments and because of horizontal gene transfer which both of them have high rates for each other. most resistant drug was penicillin’s and least resistance were carbapenems, males had higher resistance percentages than females.

Pseudomonas aeruginosa was highly resistant to all antibiotics which is concerning the reasons for these high rates are due to its intrinsic resistance factor one of which is its biofilms giving it the ability to stick to surfaces, environment exposure, limited treatment options(37), males were more resistant than females, most resistance was to gen three cephalosporins(95.7%) followed by penicillin’s(91.3%), gentamicin(82.6%), nitrofurantoin(78.3%), amikacin(73.9%) and ciprofloxacin(65.2%) these findings were also similar in (32, 33, 38, 39), the increased resistance to carbapenems(23.9%) which is caused by pseudomonas intrinsic resistance is of concern since carbapenems are reported to be the most effective and last line antibiotics(40).

Finding antimicrobial resistance trend is useful as it shows us which pathogen is the most dangerous and should be the focus of mitigation in the local community, it gives us the ability to prescribe the best antibiotic for a specific pathogen leading to better patient outcome and limiting complications, decreasing misuse and overuse of antibiotics, and also for making public guidelines on how to avoid most common pathogens to limit infections, all these points if followed will decrease resistance the local community. This research set out to find AMR trend in duhok province, we recommend new guidelines to be set for public and doctors on how misuse and overuse of antibiotics increases resistance and why antibiotics shouldn’t be used without a doctors prescription which leads to increase of resistance. This research alone isn’t enough to draw conclusions for certain and research on the subject is limited in duhok province as such we hope more research is done on AMR as it is a serious world problem and needs to be dealt with on a local community basis.

## Data Availability

All data produced in the present study are available upon reasonable request to the authors

